# Clinical Characteristics and Laboratory Biomarkers in ICU-admitted Septic Patients with and without Bacteremia: A Predictive Analysis

**DOI:** 10.1101/2023.11.16.23298625

**Authors:** Sangwon Baek, Seung Jun Lee

**Affiliations:** Department of Laboratory Medicine, Gyeongsang National University Changwon Hospital, Changwon, Republic of Korea; Center for Data Science, New York University, New York, The United States

**Keywords:** bacteremia, sepsis, prediction, procalcitonin, laboratory biomarkers, clinical biomarkers

## Abstract

**Background:** Few studies have investigated the diagnostic utilities of biomarkers for predicting bacteremia among septic patients admitted to intensive care units (ICU). Therefore, this study evaluated the prediction power of laboratory biomarkers to utilize those markers with high performance to optimize the predictive model for bacteremia.

**Methods:** A retrospective cross-sectional study was conducted at the ICU department of Gyeongsang National University Changwon Hospital in 2019. Adult patients qualifying SEPSIS-3 (increase in sequential organ failure score ≥ 2) criteria with at least two sets of blood culture were selected. Collected data was initially analyzed independently to identify the significant predictors, which was then used to build the multivariable logistic regression (MLR) model.

**Results:** A total of 218 patients with 48 cases of true bacteremia were analyzed in this research. Both CRP and PCT showed a substantial area under the curve (AUC) value for discriminating bacteremia among septic patients (0.757 and 0.845, respectively). To further enhance the predictive accuracy, we combined PCT, bilirubin, neutrophil-lymphocyte ratio (NLR), platelets, lactic acid, erythrocyte sedimentation rate (ESR), and Glasgow Coma Scale (GCS) score to build the predictive model with an AUC of 0.907 [0.843–0.956]. In addition, a high association between bacteremia and mortality rate was discovered through the survival analysis (*P*=0.004).

**Conclusions:** While PCT is certainly a useful index for distinguishing patients with and without bacteremia by itself, our MLR model indicates that the accuracy of bacteremia prediction substantially improves by the combined use of PCT, bilirubin, NLR, platelets, lactic acid, ESR, and GCS score.

## Introduction

Sepsis—a multifaceted host response to an infested pathogen—is a life-threatening disease that impairs the body’s organs and tissues [1]. The aftermath of sepsis can include multiple organ failure or septic shock that many septic patients pass away soon after the onset. According to the Surviving Sepsis Campaign, sepsis is one of the major health concerns that impacts millions of worldwide patients and constitutes approximately 15% to 30% of global deaths annually; it is also reported that sepsis is specifically dangerous among critically ill patients hospitalized in intensive care units (ICU) [2]. In addition, several reports illustrate that the mortality of septic patients particularly rises with the onset of bacteremia—a growth of pathogenic bacteria in the bloodstream [3]. Responding to these devastating reports, many researchers have conducted diverse studies on sepsis and bacteremia to improve the management strategies, and they found that early and customized therapy can substantially reduce mortality [2, 4]. To enable such a therapy, the current researchers endeavor to discover fast and precise diagnostic techniques.

Many physicians nowadays use blood culture testing results along with some established inflammatory biomarkers to diagnose sepsis and bacteremia. Though a blood culture test is an accepted assay with a reliable accuracy, it usually takes several days to detect the bacteria that this test is unsuitable for the patients requiring immediate medical treatment [5, 6]. In contrast, the diagnosis through the use of laboratory biomarkers takes only a few hours that this methodology significantly saves time. Therefore, worldwide researchers have conducted diverse studies on inflammatory biomarkers such as c-reactive protein (CRP), procalcitonin (PCT), presepsin, or interleukin-6 (IL-6) to verify their diagnostic values [7, 8]. While these biomarkers revealed an adequate accuracy in diagnosing sepsis or bacteremia, medical staffs still come to a consensus that inflammatory biomarkers are still insufficient to be used as the deterministic factor [1]. Nonetheless, there rarely has been researches on diagnostic values of clinical biomarkers, resulting the diagnostic utilities of these other biomarkers to remain unclear. Hence, this research aims to construct the new prediction model using combined laboratory biomarkers for diagnosing bacteremia among septic patients. Not limited to analyzing only CRP or PCT, we evaluated any biomarkers with remarkable diagnostic performance to utilize them for optimizing the model’s prediction power.

## Methods

### Study Setting and Design

This research was a retrospective cross-sectional study conducted at the Gyeongsang National University at Changwon Hospital (GNUCH). All hospitalized septic patients admitted to ICU from January 2019 to December 2019 were included in the study. Of 218 patients enrolled for the study, 48 were true bacteremia and 170 were non-bacteremia. All clinical data were obtained from the electronic medical record (EMR). Each patient’s information was managed through a randomly designated serial number specifically produced for this research to ensure the protection of every participant’s privacy information. The study’s informed consent and protocol were waived and approved, respectively, by the Institutional Review Board of the GNUCH (GNUCH 2021-10-010).

### Definitions

A blood culture test result was the gold standard for distinguishing patients with true bacteremia or non-bacteremia. Patients with negative culture results were defined as non-bacteremia. For patients with positive culture results, those with pathogenic bacterium were defined as true bacteremia and those with skin normal flora were reclassified to non-bacteremia. Adhering to the definition of SEPSIS-3 for determining septic status, patients with suspected infection and an acute increase in sequential organ failure assessment (SOFA) score ≥ 2 points were defined as the septic patients [9].

### Patient Enrollment

The initial candidates for the research were all patients who held the blood culture testing in 2019. The patient selection was based on the inclusion criteria predetermined at the research design to qualify only the appropriate research subjects. In case of the data duplicates per patient, the data of earlier dates were used for the analysis. The inclusion criterion for our research are as followed: 1) an ICU-admitted adult patient (Age ≥ 18); 2) a patient qualifying the criteria of SEPSIS-3 definition; 3) a patient with complete laboratory biomarkers; 4) the blood collection time difference between laboratory test and blood culture test less than ±12 hours; 5) a patient with more than two sets of both aerobic and anaerobic blood culture bottles; 6) a patient with blood collected from the peripheral vein; 7) a patient with the blood culture bottle volume data. After applying these inclusion criteria to extract the patient cohort, the medical doctor evaluated the identified bacteria of each positive culture result to verify the existence of skin normal flora. If so, those were reclassified as non-bacteremia.

### Data Collection

EMR of all selected patients was investigated to gather their baseline characteristics such as age, gender, body temperature, and laboratory results relevant to bacteremia. Our primary objective during the data collection was to accurately calculate the clinical scores while capturing the most relevant patient condition at the time of the blood culture test. Hence, the collected laboratory results represent the worst condition of the patient within ± 12 hours of the blood collection time, applying a tighter range of collection time than the data collection guidelines set by SOFA score and acute physiology and chronic health evaluation (APACHE II) score [9]. When the results of the data were questionable, that data was replaced with its comparable data or applied the formula to find its estimates. For instance, because many of our patients were intubated, their verbal Glasgow Coma Scale (GCS) subscore was calculated using motor and eye GCS subscores using the estimating formula proposed by Cheng et al. [10]. If there were no possible ways to replace the errors in data, that data was excluded. Lastly, the medical doctor documented the patients’ suspected source of infection and medical history to identify the existence of any potential confounding trend among the research subjects that may impact the study results.

### Statistical Analysis

Kolmogorov-Smirnov test was performed to check the normality of the distribution for each laboratory result. Then, a comparative analysis on laboratory biomarkers was conducted by implementing the Mann-Whiney U test for continuous variables and the chi-squared test for categorical variables. Using significant variables from the comparative analysis, we conducted the univariate logistic regression analysis to evaluate their predictive power for bacteremia. After, the multivariable logistic regression (MLR) model was constructed through a backward elimination method, in which we selected the model with the lowest Akaike’s Information Criteria and Bayesian Information Criteria [11]. Through bootstrapping 10,000 times, 95% confidence interval (CI) for sensitivity, specificity, and area under the curve (AUC) were calculated. Lastly, a survival analysis of patients with and without bacteremia using Kaplan-Meier curve analysis and a log-rank test were performed [12]. The significance level for this research is set as *P* < 0.05. All statistical analyses were performed using Python programming language (Python Software Foundation, version 3.8) and MedCalc Statistical Software version 20.015 (MedCalc Software Ltd, Ostend, Belgium).

## Results

### Clinical Characteristics

A total of 218 septic patients (increase in SOFA score ≥ 2) were included in the analysis (Fig 1). Among them, 48 (22%) were bacteremic patients and 170 (78%) were non-bacteremic patients. The median [interquartile range] age was 68 [56–77] years and males were predominant (64.2%) in the cohort (Table 1). There was no noticeable difference in age or gender proportion among patients with and without bacteremia. However, bacteremic patients had a higher proportion of in-hospital mortality than non-bacteremic patients (43.8% vs. 27.1%, *P*=0.042).

**Fig 1.**
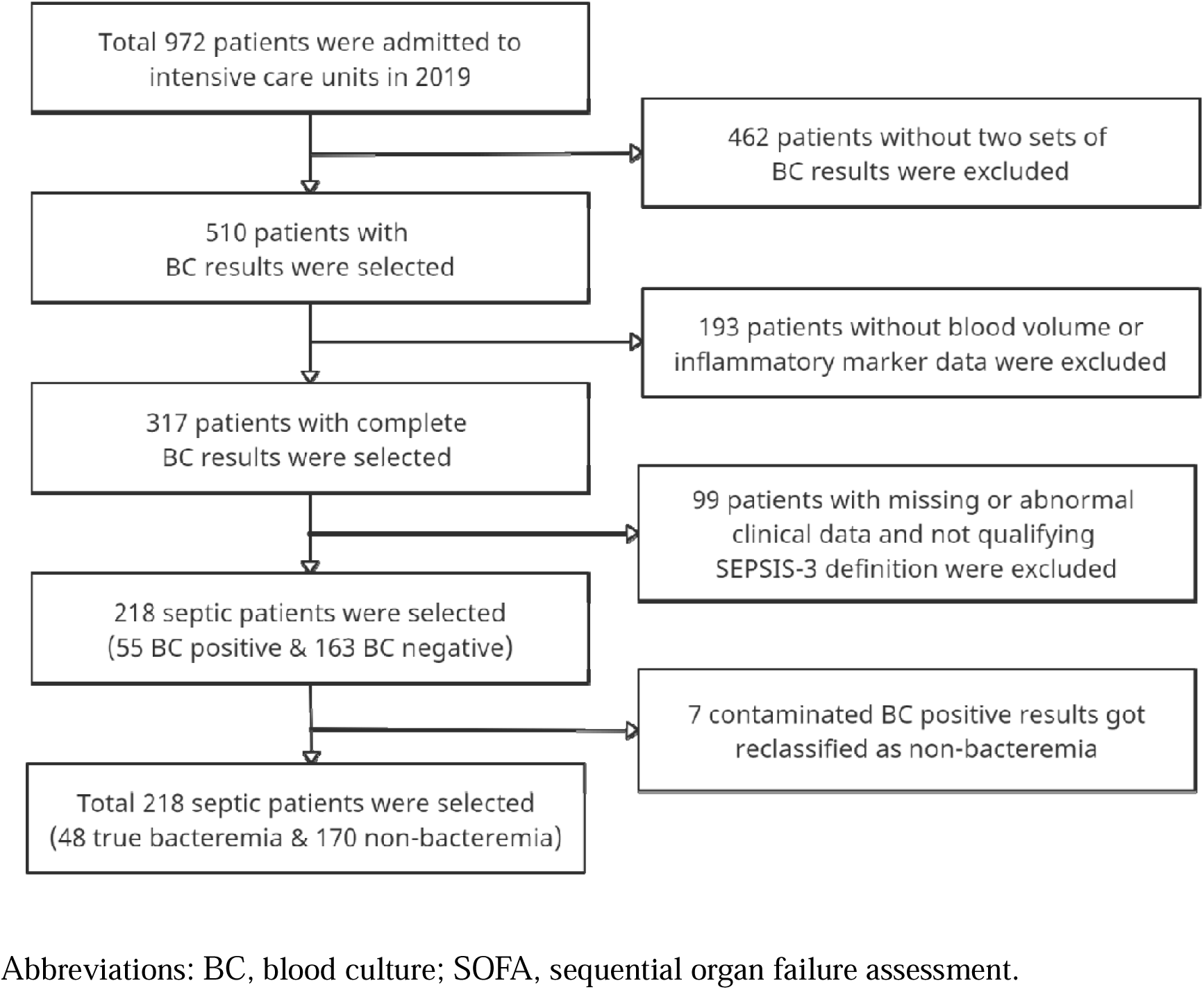
Patient enrollment flowchart

**Table 1.** Baseline characteristics of patients with and without bacteremia.

|  | Total patients<br>(N=218) | True bacteremia<br>(N=48) | Non-bacteremia<br>(N=170) | *P* |
| --- | --- | --- | --- | --- |
| **Patient Characteristics** |  |  |  |  |
| Age (years) | 68 [56–77] | 65 [55–76] | 68 [57–78] | 0.529 |
| Male sex, n (%) | 140 [64.2%] | 33 [68.8%] | 107 [62.9%] | 0.568 |
| Body temperature (°C) | 36.6 [36.3–37.1] | 37.0 [36.2–37.4] | 36.6 [36.3–37.0] | 0.066 |
| 28-day mortality (%) | 67 [30.7%] | 21 [43.8%] | 46 [27.1%] | 0.042 |
| Prior antibiotics (%) | 78 [35.8%] | 14 [29.2%] | 64 [37.6%] | 0.362 |
| **Laboratory Findings** |  |  |  |  |
| Blood culture bottle volume (ml) | 5.9 [4.8–7.6] | 5.8 [5.1–7.6] | 5.9 [4.6–7.6] | 0.455 |
| PaO<sub>2</sub>/FiO<sub>2</sub> (mmHg) | 210 [135–334] | 229 [134–353] | 208 [135–317] | 0.670 |
| Mean arterial pressure (mmHg) | 68 [60–79] | 62 [57–71] | 70 [62–79] | 0.001 |
| Serum sodium (mmol/L) | 136.4 [133.2–139.8] | 133.8 [129.0–137.3] | 137.0 [134.2–140.3] | 0.001 |
| GCS score (3–15) | 6 [3–13] | 4 [3–9] | 7 [3–14] | 0.016 |
| Neutrophil-lymphocyte ratio | 10.0 [4.7–17.8] | 18.2 [8.3–36.6] | 8.7 [4.3–14.8] | <0.001 |
| Creatinine (mg/L) | 1.04 [0.68–1.89] | 1.94 [1.02–3.06] | 0.96 [0.67–1.57] | <0.001 |
| Lactic acid (mmol/L) | 2.2 [1.1–5.6] | 3.2 [1.9–9.4] | 1.9 [1.1–3.9] | 0.001 |
| Bilirubin (mg/dL) | 0.86 [0.56–1.42] | 1.40 [0.85–2.10] | 0.77 [0.50–1.18] | <0.001 |
| Aspartic aminotransferase (U/L) | 48.0 [31.5–110.0] | 85.0 [39.5–206.0] | 44.0 [28.5–87.0] | 0.001 |
| Platelets (10<sup>3</sup>/ml) | 183 [103–265] | 111 [69–171] | 203 [127–276] | <0.001 |
| Blood urea nitrogen (mg/dL) | 22.9 [16.5–37.4] | 35.0 [23.4–56.0] | 19.9 [14.8–33.3] | <0.001 |
| White blood cell (cells/mm<sup>3</sup>) | 12.5 [7.8–18.1] | 13.4 [6.3–20.8] | 12.5 [8.0–18.1] | 0.865 |
| Hematocrit (%) | 33 [29–38] | 34 [29–36] | 33 [29–38] | 0.453 |
| Erythrocyte sedimentation rate (mm/hr) | 27 [9–53] | 43 [18–59] | 26 [7–51] | 0.039 |
| C-reactive protein (mg/L) | 86.8 [17.0–170.6] | 192.1 [90.4–259.5] | 67.9 [14.4–139.9] | <0.001 |
| Procalcitonin (ng/ml) | 0.37 [0.08–3.68] | 14.31 [3.00–45.51] | 0.22 [0.04–1.09] | <0.001 |
| Pitt bacteremia score (0–14) | 4 [2–6] | 6 [3–6] | 4 [2–6] | 0.165 |
| APACHE II score (0-71) | 20 [15–25] | 25 [18–30] | 19 [15–24] | <0.001 |
| SOFA score (0-24) | 10 [7–12] | 12 [9–16] | 9 [6–11] | <0.001 |
| **Medical History** |  |  |  |  |
| Diabetes mellitus (%) | 71 [32.6%] | 17 [35.4%] | 54 [31.8%] | 0.762 |
| Hypertension (%) | 107 [49.1%] | 22 [45.8%] | 85 [50.0%] | 0.729 |
| Heart failure (%) | 29 [13.3%] | 3 [6.2%] | 26 [15.3%] | 0.165 |
| Cerebrovascular disease (%) | 34 [15.6%] | 7 [14.6%] | 27 [15.9%] | 0.995 |
| Renal disease (%) | 27 [12.4%] | 5 [10.4%] | 22 [12.9%] | 0.825 |
| Liver disease (%) | 14 [6.4%] | 6 [12.5%] | 8 [4.7%] | 0.107 |
| Chronic obstructive pulmonary disease (%) | 16 [7.3%] | 2 [4.2%] | 14 [8.2%] | 0.521 |
| Known neoplasm (%) | 24 [11.0%] | 8 [16.7%] | 16 [9.4%] | 0.247 |
| **Suspected Source of Infection** |  |  |  |  |
| Catheter-related bloodstream infection (%) | 4 [1.8%] | 1 [2.1%] | 3 [1.8%] | >0.999 |
| Intra-abdominal infection (%) | 17 [7.8%] | 9 [18.8%] | 8 [4.7%] | 0.004 |
| Respiratory tract infection (%) | 155 [71.1%] | 21 [43.8%] | 134 [78.8%] | 0.003 |
| Skin and soft tissue infection (%) | 8 [3.7%] | 2 [4.2%] | 6 [3.5%] | >0.999 |
| Urinary tract infection (%) | 33 [15.1%] | 17 [35.4%] | 16 [9.4%] | <0.001 |
| Others (%) | 9 [4.1%] | 6 [12.5%] | 3 [1.8%] | 0.004 |
| Fever of unknown origin (%) | 6 [2.8%] | 1 [2.1%] | 5 [2.9%] | >0.999 |

Continuous variables are expressed in median [interquartile range] and categorical variables are expressed in counts [proportions (%)]. Mann–
Whitney U test performed for continuous variables and chi-squared contingency test or Fisher's exact test, where appropriately, were performed for categorical variables to calculate *P*.
Abbreviations: GCS, Glasgow coma scale; APACHE II, acute physiology and chronic health evaluation II; SOFA, sequential organ failure assessment.

The clinical profiling between patients with and without bacteremia were significantly different. Those with bacteremia had substantially lower mean arterial pressure (MAP, 62 [57–71] vs. 70 [62–79]), lower serum sodium (Na) level (133.8 [129.0–137.3] vs. 137.0 [134.2–140.3]), lower GCS score (4 [3–9] vs. 7 [3–14]), and lower platelet counts (111 [69–171] vs. 203 [127–276). In addition, bacteremic patients had substantially higher neutrophil-lymphocyte ratio (NLR, 18.2 [8.3–36.6] vs. 8.7 [4.3–14.8]), higher creatinine (1.96 [1.02–3.06] vs. 0.96 [0.67–1.57]), higher lactic acid (3.2 [1.9–9.4] vs. 1.9 [1.1–3.9]), higher bilirubin (1.40 [0.85–2.10] vs. 0.77 [0.50–1.18]), higher aspartic aminotransferase (AST, 85.0 [39.5–206.0] vs. 44.0 [28.5–87.0]), higher blood urea nitrogen (BUN, 35.0 [23.4–56.0] vs. 19.9 [14.8–33.3]), higher erythrocyte sedimentation rate (ESR, 43 [18–59] vs. 26 [7–51]), higher CRP (192.1 [90.4–259.5] vs. 67.9 [14.4–139.9]), and higher PCT (14.31 [3.00–45.51] vs. 0.22 [0.04–1.09]). Bacteremia severity scores, such as APACHE II (25 [18–30] vs. 19 [15–24]) and SOFA (12 [9–16] vs. 9 [6–11]), were higher in bacteremic patients. Last but not least, bacteremic patients were more likely to be infected through an intra-abdominal infection (18.8% vs. 4.7%) and an urinary tract infection (35.4% vs. 9.4%). Whereas a higher proportion of non-bacteremic patients were infected through a respiratory tract infection (78.8% vs. 43.8%).

### Predictive Value of Clinical Markers

The significant variables identified through the comparative analysis also showed statistical significance in a univariate logistic regression analysis (Table 2). Among them, PCT (AUC, 0.845 [0.771–0.907]; DOR, 21.29) and CRP (AUC, 0.757 [0.670–0.833]; DOR, 8.45) had the highest diagnostic performance with optimal cutoff values of 3.18 ng/ml and 164.3 mg/L, respectively. Both CRP and PCT even had the higher predictive power than SOFA score—the aggregate of clinical indices that is even used as the SEPSIS-3 diagnosis criteria nowadays [1]. In addition, a subgroup analysis of gram-positive bacteria (GPB) vs. gram-negative bacteria (GNB) revealed that the patients with GNB bacteria are likely to have substantially higher PCT concentration than the patients with GPB bacteria—in line with the previous findings (See Supplemental Data Table S1) [13].

**Table 2.** Univariate and multivariable logistic regression analysis to predict bacteremia using the significant biomarkers.

|  | **Cut-off** | **AUC** | **Sensitivity** | **Specificity** | **PPV** | **NPV** | **LR +** | **LR -** | **DOR** | **P** |
| --- | --- | --- | --- | --- | --- | --- | --- | --- | --- | --- |
| **MLR model\*** |  | 0.907 [0.843–0.956] | 87.9% [74.5–96.6] | 86.6% [78.8–96.0] | 59.7% | 96.6% | 5.25 | 0.13 | 41.81 | <0.001 |
| **PCT (ng/ml)** | 3.18 | 0.845 [0.771–0.907] | 77.3% [64.4–88.9] | 87.2% [79.1–92.5] | 63.2% | 92.5% | 6.07 | 0.29 | 21.29 | <0.001 |
| **CRP (mg/L)** | 164.3 | 0.757 [0.670–0.833] | 63.8% [48.8–84.2] | 84.2% [59.5–90.9] | 51.7% | 88.8% | 3.79 | 0.45 | 8.45 | <0.001 |
| **SOFA score (0-24)** | 13 | 0.734 [0.650–0.814] | 60.8% [38.5–92.2] | 80.4% [41.3–93.2] | 50.0% | 85.5% | 3.54 | 0.6 | 5.88 | <0.001 |
| **Bilirubin (mg/dL)** | 0.97 | 0.733 [0.647–0.809] | 70.9% [42.1–91.1] | 69.1% [45.3–93.2] | 37.2% | 89.5% | 2.1 | 0.41 | 5.07 | <0.001 |
| **BUN (mg/dL)** | 19.2 | 0.712 [0.624–0.791] | 82.7% [47.3–96.0] | 57.4% [41.5–89.9] | 31.9% | 94.0% | 1.66 | 0.23 | 7.29 | <0.001 |
| **NLR** | 14.26 | 0.709 [0.612–0.798] | 65.9% [46.7–80.4] | 76.6% [66.1–88.4] | 41.0% | 88.6% | 2.46 | 0.46 | 5.39 | <0.001 |
| **Platelets (10<sup>3</sup>/ml)** | 168 | 0.707 [0.616–0.793] | 77.8% [56.2–91.3] | 63.5% [50.3–80.1] | 36.4% | 89.9% | 2.02 | 0.4 | 5.1 | <0.001 |
| **Creatinine (mg/L)** | 1.86 | 0.702 [0.619–0.780] | 61.0% [46.2–88.5] | 81.8% [50.9–88.3] | 48.2% | 87.0% | 3.3 | 0.53 | 6.25 | 0.005 |
| **APACHE II score (0-71)** | 24 | 0.673 [0.573–0.766] | 56.2% [34.0–79.5] | 78.6% [51.7–93.1] | 37.8% | 86.1% | 2.16 | 0.57 | 3.77 | <0.001 |
| **Na (mmol/L)** | 133.4 | 0.662 [0.568–0.750] | 54.8% [32.6–87.0] | 79.2% [42.4–93.3] | 40.7% | 84.9% | 2.43 | 0.63 | 3.86 | 0.004 |
| **Lactic acid (mmol/L)** | 6.1 | 0.657 [0.564–0.741] | 64.6% [34.6–91.8] | 64.3% [33.7–90.0] | 41.7% | 83.5% | 2.53 | 0.7 | 3.62 | 0.008 |
| **AST (U/L)** | 62 | 0.655 [0.562–0.742] | 65.0% [41.7–89.8] | 67.2% [36.0–84.8] | 34.1% | 86.6% | 1.83 | 0.55 | 3.34 | 0.015 |
| **MAP (mmHg)** | 67.3 | 0.653 [0.562–0.742] | 68.4% [42.4–87.3] | 63.1% [41.1–83.0] | 32.4% | 87.1% | 1.69 | 0.53 | 3.22 | 0.003 |
| **GCS score (3-15)** | 10 | 0.611 [0.526–0.694] | 79.6% [50.0–93.8] | 43.8% [25.4–70.6] | 27.5% | 88.2% | 1.34 | 0.48 | 2.82 | 0.017 |
| **ESR (mm/hr)** | 41 | 0.597 [0.503–0.690] | 59.6% [29.4–89.2] | 67.4% [31.8–90.3] | 32.9% | 84.6% | 1.74 | 0.65 | 2.69 | 0.029 |

Cut-off is calculated through using the Youden's index and 95% confidence interval is calculated through bootstrapping the logistic regression model 10,000 times.
\*This index is constructed by combining PCT, Bilirubin, NLR, Platelets, Lactic acid, GCS score, and ESR.
Abbreviations: PPV, positive predictive value; NPV, negative predictive value; LR+, positive likelihood ratio; LR-, negative likelihood ratio; DOR, diagnostic odds ratio; CI, confidence interval; AUC, area under the curve; MLR, multivariable logistic regression; PCT, procalcitonin; CRP, c-reactive protein; SOFA, sequential organ failure assessment; BUN, blood urea nitrogen; NLR, neutrophil-lymphocyte ratio; APACHE II, acute physiology and chronic health evaluation II; Na, serum sodium; AST, aspartic aminotransferase; MAP, mean arterial pressure; GCS, Glasgow coma scale; ESR, erythrocyte sedimentation rate.

After performing the univariate analysis, we then further enhanced the diagnostic model for differentiating patients with and without bacteremia through constructing the MLR model. This MLR model consisted of PCT, bilirubin, NLR, platelets, lactic acid, ESR, and GCS score as the predictors, and it had the prediction power with an AUC of 0.907 [0.843– 0.956] (See Supplemental Data Table S2). The degree of improvement in diagnostic performance was visually represented through ROC curves where each variable’s predictive accuracy was compared to that of the MLR model (Fig 2). A noticeable insight from the ROC plot is that the prediction power has increased dramatically for a multivariable model compared with univariate models. Last but not least, the difference in survival probability of septic patients with and without bacteremia is shown through the Kaplan-Meier curve (Fig 3). The plot demonstrates that septic patients without bacteremia were 20% more likely to survive compared to those with bacteremia.

**Fig 2.**
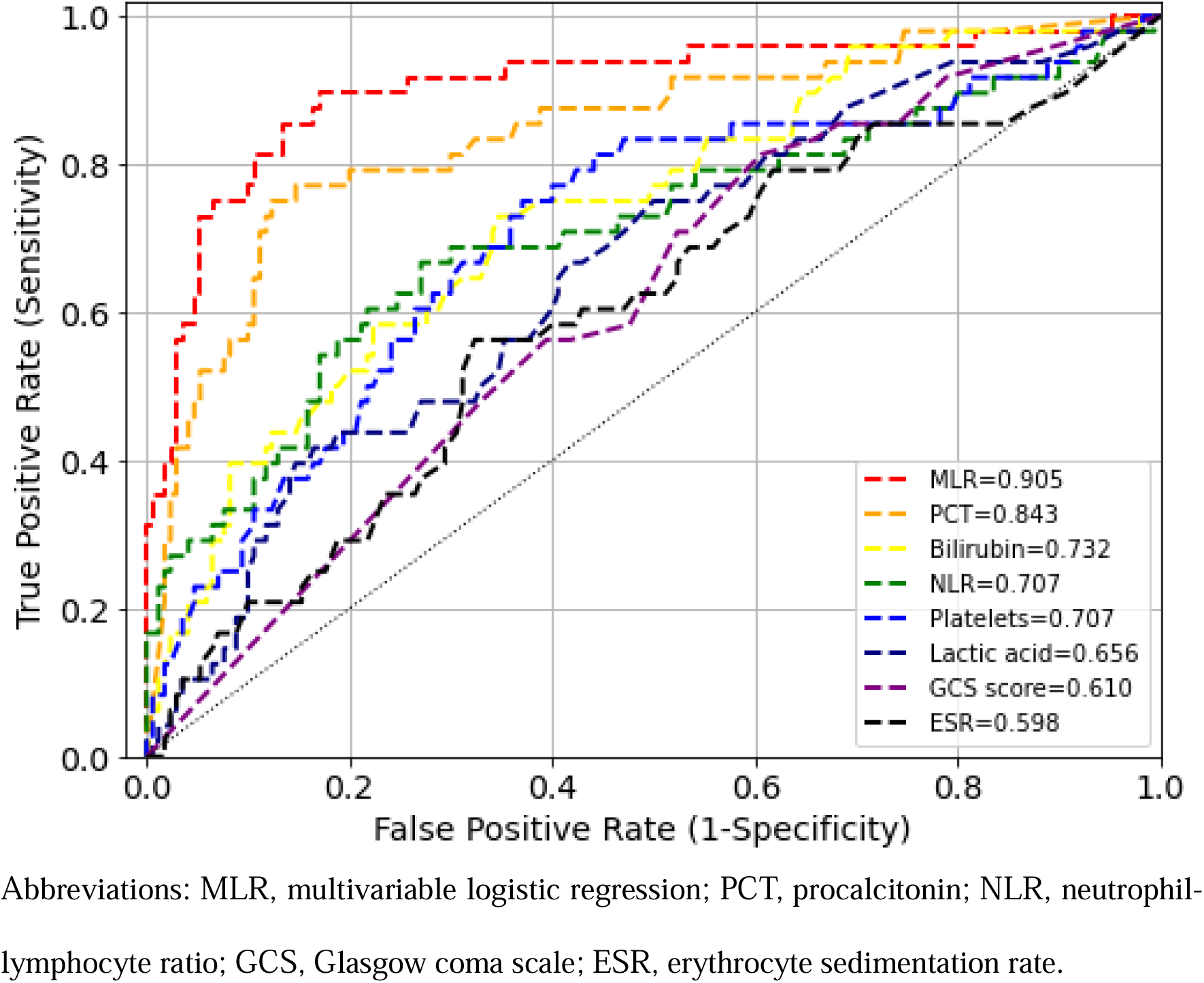
Plot of ROC curves of univariate and multivariable prediction model for bacteremia

**Fig 3.**
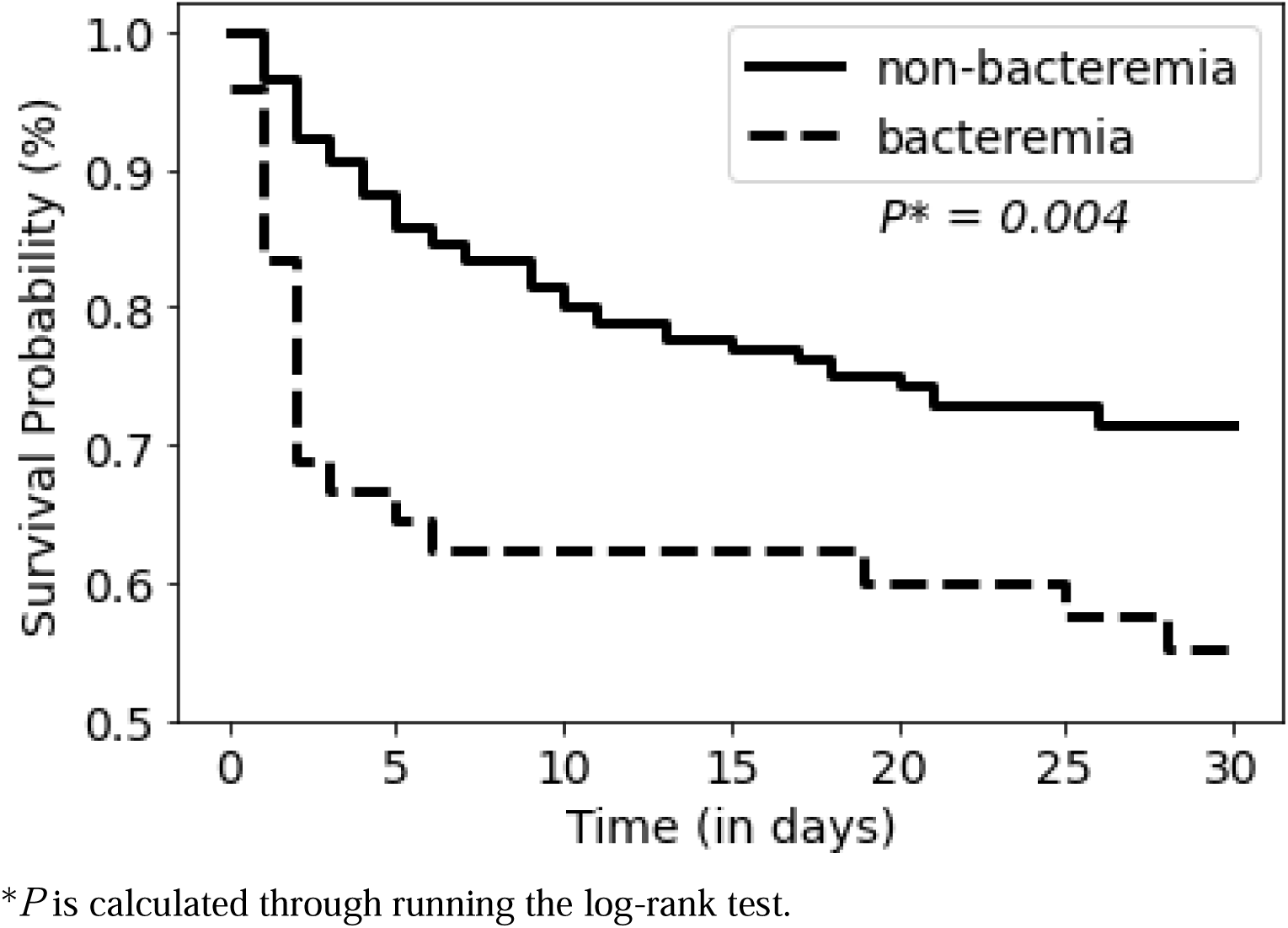
Plot of Kaplan-Meier estimates for the survival of bacteremic vs. non-bacteremic patients

## Discussion

### Diagnostic Utilities of CRP and PCT

Inflammatory biomarkers have been acknowledged as valuable predictors for sepsis and bacteremia that many previous researchers have investigated to confirm their diagnostic utilities [14–20]. Of all inflammatory biomarkers, the popular indices known for their association with bacteremia and sepsis are CRP, PCT, presepsin, and IL-6. Many studies have used these markers to build the prediction model for bacteremia and sepsis [8, 21, 22]. One meta-analysis reported the diagnostic efficiency of presepsin particularly during early-stage sepsis because presepsin level increases prior to CRP and PCT [23]. The same study also argued that presepsin could be useful for increasing the accuracy of the prediction model when used in combination with other markers such as PCT or IL-6 [23]. Although analyzing presepsin and IL-6 could increase the accuracy of the model, this research primarily focused on analyzing CRP and PCT due to the lack of presepsin and IL-6 data. The reported predictive performance from previous studies ranged between AUC of 0.56–0.66 for CRP and AUC of 0.68–0.79 for PCT [15, 17, 22]. In contrast to these results, our research showed a significantly higher predictive accuracy with an AUC of 0.757 for CRP and 0.845 for PCT. A possible reason for such difference in prediction power attributes to the diverse study population that the baseline patient conditions could have been a confounding factor. Meanwhile, our study demonstrated that PCT is a better predictor for bacteremia than CRP, which was in line with previous studies. From the clinical standpoint, such distinction is reasonable because CRP—an acute-phase protein—often rises due to a wide variety of conditions such as inflammation, infection, and tissue damage, while PCT—a peptide precursor of calcitonin—usually rises specifically in response to bacterial infection [22, 24].

### Diagnostic Utilities of Biomarkers included in MLR Model

Through the MLR modeling, we further enhanced the prediction power using PCT, bilirubin, NLR, platelets, lactic acid, ESR, and GCS score. Although CRP and SOFA showed a high prediction power in the univariate analysis, these variables were eliminated from the MLR model due to their collinearity with other included biomarkers. The combination of the variable constituting the MLR model is unconventional, yet the majority of these variables show a high correlation with bacteremia. Bilirubin—an orange-yellow substance that forms in response to break down of red blood cells—is a well-known index for displaying the liver condition. According to Yan et al., the liver has multiple functions that include clearing out the pathogenic bacteria in the body system; therefore, hyperbilirubinemia—indicative of a liver dysfunction—exposes the patients to a higher risk of developing bacteremia [25]. NLR—a known marker for bacteremia—indicates the disease severity because neutrophil counts increase and lymphocyte counts decrease consistently with inflammation progress [26]. A recent study highlighted NLR as the significant predictor for bacteremia with an AUC of 0.83 [0.66–0.80] [27]. A decrease in platelet—tiny blood cells that control bleeding in a body—counts often lead to coagulation abnormality. One research on critically ill patients demonstrated the high association between thrombocytopenia and bacteremia, for which the researchers emphasized the need of using platelet counts for better evaluations of patients with bacterial infection [28]. A study held by the emergency department evaluated the diagnostic value of lactic acid by combining it with CRP and PCT. In line with our study result, this study also discovered a high diagnostic utility of lactic acid where the diagnostic odds ratio for lactic acid alone was 2.93 [2.09–4.14] and lactic acid+PCT was 3.98 [2.81– 5.63] [29]. ESR and GCS score showed the least correlation with bacteremia among included biomarkers, which indicate trivial utilities of ESR and GCS score as independent variables. However, the prediction power of our MLR model was enhanced by including ESR and GCS score that these variables may have significant predictive utilities when used in combination with other indices.

### Comparative Analysis on Discriminatory Ability of the Prediction Models

We constructed the optimal prediction for bacteremia with an AUC of 0.907 [0.843– 0.956]. There have been several attempts to optimize the prediction accuracy by combining diverse independent variables like that of our model. One study constructed the prediction model for bacteremia (AUC, 0.851 [0.828–0.875]) among 2,888 cirrhotic patients by combining bilirubin, albumin, WBC, platelets, CRP, and creatinine [30]. Another study constructed the prediction model (AUC, 0.70 [0.67–0.73]) for bacteremia among septic patients qualifying SEPSIS-3 by combining PCT, CRP, Lactic acid, and NLR. As shown, our study discovered predictor combinations different from conventional preexisting models, and our model also outperformed the prediction power among existing multivariable models. Though an inquiry into the combinations of biomarkers is necessary to enhance the prediction power, our model is certainly a pioneering step towards diagnostic advancement.

### Clinical Outcomes of Septic Patients With and Without Bacteremia

Our prediction model is also beneficial from the clinical perspective because our study revealed a high association between bacteremia and in-hospital mortality. The Kaplan-Meier curve demonstrated that the septic patients without bacteremia were 20% more likely to survive than those with bacteremia during their 28-day hospitalization, and a log-rank test with *P*=0.004 confirmed that such difference in survival probability is statistically significant (Fig 3). In contrast, previous studies have found diverse results for mortality rates in bacteremic and non-bacteremic patients that some reported a strong association while others reported no significance [3, 8, 13, 14, 16]. One possible reason for such conflicting results may attribute to different study designs. For instance, whether appropriate antibiotics were administered before the blood culture test or not can significantly influence the culture result. Also, the study population had diverse baseline patient characteristics that the clinical manifestation may have been different according to their pre-existing condition. Despite the varying reports on mortality rate which may require further research, our study has shown a strong correlation between bacteremia and mortality among our selected cohort; such outcome serves to signify the clinical value of determining septic patients with bacteremia through our prediction model.

### Limitations

The limitations of the study must be addressed. First, the baseline characteristics of our patient cohorts were unusual. Most non-bacteremic patients had respiratory tract infections and most bacteremic patients had either intra-abdominal infections or urinary tract infections. There existed a statistically significant distinction in the source of infection that could have been a confounding factor. Second, this was a retrospective study that excluding patients with missing or abnormal data was inevitable in our research setting. Such patient exclusion may have led to the selection bias in our research subjects. Third, the patients with insufficient blood volume for blood culture testing were included. Although collecting 8–12 ml of volume can substantially reduce the contamination rates of blood culture, such volume range could not be set as the inclusion criteria due to the lack of sample size [5]. Fortunately, the blood volume for patients with and without bacteremia did not show a significant difference (*P*=0.45) that we expected the same effect on both cohorts would nullify its influence. Fourth, we were unable to follow up with the patients that their outcome after the discharge from our hospital is unknown. Finally, the patients with administered antibiotics were included in the analysis to retain the sample size. Though antibiotics can affect the blood culture result, there was no significant difference in proportions among bacteremic and non-bacteremic patient cohorts (*P*=0.36) that we expect the influence to be mitigated.

### Conclusions

In summary, we constructed the optimal prediction model for bacteremia through MLR modeling. Also, our research showed the strong association between bacteremic sepsis and mortality that this model has significant clinical utilities for enabling rapid and precise diagnosis among patients with and without bacteremia. Although more research seems necessary to confirm the applicability or validity of our new prediction model, we confirmed that PCT alone has adequate diagnostic value and its prediction power could be maximized by adding bilirubin, NLR, platelets, lactic acid, ESR, and GCS score.

## Data Availability

The datasets used and/or analyzed during the current available from the corresponding author on reasonable request.

## Acknowledgments

The authors thank GNUCH’s physicians working at the emergency department and the ICU department for providing valuable data for this study.

## Author Contributions

Baek SW designed the study, analyzed the data, interpreted the results, and wrote the manuscript. Lee SJ supervised the study design, evaluated the clinical data, and revised the manuscript. All authors read and approved the final manuscript.

## Declaration

### Ethics approval and consent to participate

Ethics approval and consent for participation Informed consent forms were obtained from all study participants. Informed consent forms were waived. This study was approved by Institutional Review Board of Gyeongsang National University Changwon Hospital (GNUCH 2021-10-010). All methods and informed consent process were performed in accordance with the relevant guidelines and regulations by Institutional Review Board of GNUCH.

### Consent for Publication

Not applicable

