## Supplementary Tables for "Clinical Characteristics and Laboratory Biomarkers in ICU-admitted Septic Patients with and without Bacteremia: A Predictive Analysis"

Table S1. Subgroup analysis of GPB vs. GNB among patients with bacteremia (n=44).

|  | **GPB (n=14)** | **GNB (n=30)** | ***P*** |
| --- | --- | --- | --- |
| **PCT (ng/ml)** | 3.00 [0.33–11.24] | 26.64 [6.01–51.10] | 0.019 |
| **CRP (mg/L)** | 146.3 [39.4–202.4] | 208.4 [97.3–259.5] | 0.137 |
| **SOFA score (0-24)** | 11 [7–13] | 12 [9–16] | 0.225 |
| **NLR** | 15.0 [5.8–37.1] | 18.6 [11.6–36.6] | 0.332 |
| **AST (U/L)** | 68.0 [37.0–101.0] | 88.5 [45.5–206.0] | 0.345 |
| **Na (mmol/L)** | 134.1 [131.9–139.1] | 132.9 [129.0–137.1] | 0.520 |
| **Platelets (10^3^/ml)** | 134 [89–175] | 106 [73–166] | 0.520 |
| **Lactic acid (mmol/L)** | 4.7 [1.9–11.8] | 3.0 [2.0–7.4] | 0.529 |
| **Creatinine (mg/L)** | 1.94 [1.12–2.38] | 1.96 [1.00–3.17] | 0.623 |
| **BUN (mg/dL)** | 35.0 [22.2–42.2] | 29.9 [23.4–56.8] | 0.668 |
| **APACHE II score (0-71)** | 23 [18–30] | 25 [20–29] | 0.733 |
| **ESR (mm/hr)** | 43 [19–68] | 42 [20–54] | 0.830 |
| **28-day mortality** | 9 [64.3%] | 17 [56.7%] | 0.881 |
| **Bilirubin (mg/dL)** | 1.48 [0.76–2.08] | 1.3 [0.95–1.96] | 0.920 |
| **MAP (mmHg)** | 63 [56–71] | 63 [57–71] | 0.940 |
| **GCS score (3-15)** | 5 [3–7] | 3 [3–9] | 0.979 |

Continuous variables are expressed in median [interquartile range] and categorical variables are expressed in counts [proportions (%)]. Mann-Whitney U test was performed for continuous variables and chi-squared contingency test was performed for categorical variables to calculate *P*.

Abbreviations: GPB, gram-positive bacteria; GNB, gram-negative bacteria; PCT, procalcitonin; CRP, c-reactive protein; SOFA, sequential organ failure assessment; NLR, neutrophil-lymphocyte ratio; AST, aspartic aminotransferase; Na, serum sodium; BUN, blood urea nitrogen; APACHE II, acute physiology and chronic health evaluation II; ESR, erythrocyte sedimentation rate; MAP, mean arterial pressure; GCS, Glasgow coma scale.

Table S2. The variables of multivariable logistic regression model

|  | **β Coefficient** | **OR (95% CI)** | ***P*** |
| --- | --- | --- | --- |
| Intercept | -2.72 |  |  |
| Procalcitonin (ng/ml) | 0.045 | 1.046 [1.022–1.071] | <0.001 |
| NLR | 0.061 | 1.063 [1.028–1.098] | <0.001 |
| ESR (mm/hr) | 0.031 | 1.031 [1.012–1.050] | 0.001 |
| Platelets (10^3^/ml) | -0.007 | 0.993 [0.988–0.998] | 0.004 |
| Lactic acid (mmol/L) | 0.109 | 1.116 [1.027–1.211] | 0.009 |
| Bilirubin (mg/dL) | 0.325 | 1.384 [1.052–1.820] | 0.02 |
| GCS score (3-15) | -0.132 | 0.876 [0.784–0.979] | 0.02 |

The independent predictors were selected through implementing backward elimination method with the significance level of 0.05.

Abbreviations: OR, odds ratio; NLR, neutrophil-lymphocyte ratio; ESR, erythrocyte sedimentation rate; GCS, Glasgow coma scale.
